# Antigen-based multiplex strategies to discriminate SARS-CoV-2 natural and vaccine induced immunity from seasonal human coronavirus humoral responses

**DOI:** 10.1101/2021.02.10.21251518

**Authors:** Eric D. Laing, Spencer L. Sterling, Stephanie A. Richard, Nusrat J. Epsi, Si’Ana Coggins, Emily C. Samuels, Shreshta Phogat, Lianying Yan, Nicole Moreno, Christian L. Coles, Matthew Drew, Jennifer Mehalko, Caroline E. English, Scott Merritt, Katrin Mende, Vincent J. Munster, Emmie de Wit, Kevin K. Chung, Eugene V. Millar, David R. Tribble, Mark P. Simons, Simon D. Pollett, Brian K. Agan, Dominic Esposito, Charlotte Lanteri, G. Travis Clifton, Edward Mitre, Timothy H. Burgess, Christopher C. Broder

**Affiliations:** Department of Microbiology and Immunology, Uniformed Services University of the Health Sciences, Bethesda, MD, USA; Henry M. Jackson Foundation for the Advancement of Military Medicine, Inc., Bethesda, MD USA; Infectious Disease Clinical Research Program, Department of Preventive Medicine and Biostatistics, Uniformed Services University of the Health Sciences, Bethesda, MD, USA; Protein Expression Laboratory, National Cancer Institute RAS Initiative, Frederick National Laboratory for Cancer Research, Frederick, MD, USA; Brooke Army Medical Center, JBSA Fort Sam Houston, TX, USA; Laboratory of Virology, Rocky Mountain Laboratories, National Institute of Allergy and Infectious Diseases, National Institutes of Health, Hamilton, MT, USA; Department of Medicine, Uniformed Services University of the Health Sciences, Bethesda, MD, USA

**Author notes:** Correspondence Eric D. Laing, PhD, Department of Microbiology and Immunology, Uniformed Services University, Bethesda, MD 20814, Christopher C. Broder, PhD, Department of Microbiology and Immunology, Uniformed Services University, Bethesda, MD 20814.

## Abstract

Sensitive and specific SARS-CoV-2 antibody assays remain critical for community and hospital-based SARS-CoV-2 sero-surveillance. With the rollout of SARS-CoV-2 vaccines, such assays must be able to distinguish vaccine from natural immunity to SARS-CoV-2 and related human coronaviruses. Here, we developed and implemented multiplex microsphere-based immunoassay strategies for COVD-19 antibody studies that incorporates spike protein trimers of SARS-CoV-2 and the endemic seasonal human coronaviruses (HCoV), enabling high throughout measurement of pre-existing cross-reactive antibodies. We varied SARS-CoV-2 antigen compositions within the multiplex assay, allowing direct comparisons of the effects of spike protein, receptor-binding domain protein (RBD) and nucleocapsid protein (NP) based SARS-CoV-2 antibody detection. Multiplex immunoassay performance characteristics are antigen-dependent, and sensitivities and specificities range 92-99% and 94-100%, respectively, for human subject samples collected as early as 7-10 days from symptom onset. SARS-CoV-2 spike and RBD had a strong correlative relationship for the detection of IgG. Correlation between detectable IgG reactive with spike and NP also had strong relationship, however, several PCR-positive and spike IgG-positive serum samples were NP IgG-negative. This spike and NP multiplex immunoassay has the potential to be useful for differentiation between vaccination and natural infection induced antibody responses. We also assessed the induction of *de novo* SARS-CoV-2 IgG cross reactions with SARS-CoV and MERS-CoV spike proteins. Furthermore, multiplex immunoassays that incorporate spike proteins of SARS-CoV-2 and HCoVs will permit investigations into the influence of HCoV antibodies on COVID-19 clinical outcomes and SARS-CoV-2 antibody durability.

## INTRODUCTION

Severe acute respiratory syndrome coronavirus-2 (SARS-CoV-2) is a novel zoonotic positive-sense, single-stranded, RNA virus responsible for the third viral pandemic of the 21^st^ century, and the third zoonotic coronavirus outbreak in the past 20 years (1, 2). At this time, SARS-CoV-2 has globally caused 106 million COVID-19 cases and over 2 million COVID-19 related deaths. Serology studies have demonstrated SARS-CoV-2 infection elicits an antibody responses that can persist for as long as 8 months (3), and that the magnitude of the antibody response is associated with COVID-19 severity (4, 5). A variety of antibody tests have been developed and granted Emergency Use Authorization (EUA) by the U.S. Food and Drug Administration (6), with the majority of these tests designed to assess for antibodies against the SARS-CoV-2 spike (S) envelope glycoprotein, the primary target of virus-neutralizing antibodies (7), in either its native-like oligomer conformation, or against one of its protein subunits or domains.

A native-like SARS-CoV-2 prefusion stabilized S-2P glycoprotein ectodomain trimer (hereafter referred to as spike) (8, 9) has been adopted for large-scale SARS-CoV-2 antigen-based serology and serosurveillance (10-13). The receptor-binding domain (RBD) located within the more variable S1 subunit of the S glycoprotein, lacking potentially conserved epitopes with endemic seasonal human coronavirus (HCoV) S glycoproteins and conferring specificity for SARS-CoV-2, has also been extensively used in antigen-based immunoassays (10, 14-16). Furthermore, immunoassay detection of IgG antibodies that can bind to RBD has been used as a surrogate for neutralization tests which require cell-culture, pseudoviruses, or high biosafety-containment and wild-type SARS-CoV-2 (15, 17, 18). Lastly, the SARS-CoV-2 nucleocapsid protein (NP) has been used in several lateral flow and antigen-based COVID-19 serology tests (6).

A handful of microsphere-based SARS-CoV-2 serology assays have been developed, facilitating high throughput multiplex strategies for antibody detection (19-23). Multiplex microsphere-based immunoassays (MMIA) have several advantages over traditional immunoassays including optical improvements in sensitivity and specificity, as well as reductions in sample volume and materials to test for antigen-specific antibodies since multiple target antibodies can be simultaneously captured with multiple antigens coupled to unique fluorescent microspheres. Additionally, multiplexing systems, e.g. Luminex xMAP-based platforms, have a large dynamic range, which have been shown to be more sensitive than ELISA for the detection of antibodies to viral infections (21, 24-26). As COVID-19 vaccine rollouts continue in the U.S., MMIA strategies allowing simultaneous detection of spike and NP reactive antibodies may facilitate differentiation of SARS-CoV-2 natural infection and vaccination (27), as all current U.S. FDA EUA COVID-19 vaccines induce humoral responses to the S glycoprotein (28, 29).

We have previously utilized a SARS-CoV-2 MMIA in cross-sectional serological analysis of military healthcare workers deployed to the Jacob K. Javits Center COVID-19 field hospital, (Javits Medical Station) (30) and U.S. Navy personnel deployed on the USNS COMFORT (31) and during the first SARS-CoV-2 epidemic wave in New York City, NY. Utilizing sera from three cohorts, a) SARS-CoV-2 naïve serum samples from the Acute Respiratory Infection Consortium Natural History Study (ARIC) collected from 2012 – 2018 (32), b) serum samples from PCR-confirmed SARS-CoV-2 subjects enrolled in the ongoing Epidemiology, Immunology, and Clinical Characteristics of Emerging Infectious Diseases with Pandemic Potential (EPICC) study and c) serum samples from hospitalized patients at the Javits Medical Station (JMS) who participated in the COVID-19 Antibody Prevalence in Military Personnel Deployed to New York (CAMP-NYC) study (30), we describe the utility of three distinct antigen-based MMIAs for COVID-19 serology studies.

Initially, we examined sensitivity differences in detection of SARS-CoV-2 antibodies between widely used antigens: SARS-CoV-2 prefusion stabilized S-2P glycoprotein ectodomain trimer (spike), a monomeric receptor-binding domain (RBD) protein and nucleoprotein (NP). Additionally, given the high seroprevalence of the seasonal HCoVs (33-35) and evidence of pre-existing antibody cross-reactivity with SARS-CoV S glycoprotein (36, 37), we validated improvements to assay specificity by the concurrent measurement of SARS-CoV-2 and HCoV-specific immunoglobulin G (IgG) in a multiplex approach and report the stimulation of cross-reactive antibody responses across ARIC, EPICC and JMS cohorts. Furthermore, we standardized SARS-CoV-2 and HCoV MMIA SARS-CoV-2 IgG detection in paired venous and capillary blood specimens collected by serum separator tubes (SST) and dried blood spots (DBS), respectively.

The objectives of this study were to develop and validate a high throughput antigen based assay which can discriminate SARS-CoV-2 from seasonal human coronavirus (HCoV) infections, as well as SARS-CoV-2 vaccination, including from specimens collected through self-collected DBS specimens which may greatly facilitate the performance of post vaccination serosurveys and vaccine effectiveness studies. This SARS-CoV-2/HCoV MMIA strategy also has the potential to enhance investigations of the interplay of pre-existing seasonal HCoV antibodies on SARS-CoV-2-specific antibody durability, COVID-19 symptom presentation and disease severity, and we present exploratory results here which show that coronavirus serological cross-reactivity is influenced by the severity of SARS-CoV-2 illness.

## MATERIALS AND METHODS

### Recombinant protein antigens and microsphere coupling

Prefusion stabilized SARS-CoV-2 spike (S) glycoprotein was used in serology testing to capture the full humoral response including all conformation-dependent antibodies. Prefusion stabilized SARS-CoV-2 S-2P glycoprotein ectodomain trimers (spike protein) and SARS-CoV-2 RBD were purchased from LakePharma, Inc. (Hopkinton, MA USA). This SARS-CoV-2 spike protein shares an equivalent ectodomain with the NIH Vaccine Research Center designed SARS-CoV-2 spike protein, and the Mount Sinai SARS-CoV-2 spike protein used in ELISA-based serology (9, 10, 16, 38, 39).

Design and expression of prefusion stabilized betacoronavirus (β-CoV) HCoV-HKU1, HCoV-OC43, SARS-CoV and MERS-CoV spike proteins have been previously described (37, 38). The alphacoronavirus (α-CoV) HCoV-229E and HCoV-NL63 spike proteins were similarly constructed and prepared (LakePharma, Inc.). The SARS-CoV-2 nucleocapsid protein (NP) was purchased from RayBiotech, Inc. (Peachtree Corners, GA, USA). A mock antigen, consisting of cell culture supernatant from non-transfected HEK cells was collected via centrifugation then filtered through a 0.22 µM PES filter to remove debris. Mock antigen-coupled beads were included in each microtiter well to control for non-specific/artificial antisera binding; samples that react with the mock antigen above an established 3-fold cutoff were retested. Proteins were coupled to carboxylated magnetic MagPlex microspheres (Bio-Rad, Hercules, CA) at a protein to microsphere ratio of 15 µg:100 µL, and antigen-coupled microspheres were resuspended in a final volume of 650 µL following manufacturer’s protocol (Bio-Rad) for amine coupling.

### Participant enrollment and sera collection

SARS-CoV-2 negative human serum specimens utilized were from sera collected from 2012 – 2018 in the ARIC Natural History Study (IDCRP-045) (32). ARIC sera predate the COVID-19 pandemic and were collected from subjects who had nasopharyngeal swabs tested by nucleic acid amplification methods for virus etiologies of acute respiratory infections; samples collected from individuals with rhinovirus and the seasonal HCoVs, HCoV-OC43, -HKU1, -229E and -NL63 were used (40). In addition, we utilized serum samples collected since the emergence of SARS-CoV-2 under the IDCRP EPICC (IDCRP-085) protocol; a prospective, longitudinal observational cohort study to analyze the natural history of COVID-19 disease. Subjects were enrolled at five hospitals across the continental U.S., including Walter Reed National Military Medical Center (WRNMMC, Bethesda, MD), Brooke Army Medical Center (BAMC, San Antonio, TX), Naval Medical Center San Diego (NMCSD, San Diego, CA), Madigan Army Medical Center (MAMC, Tacoma, WA) and Fort Belvoir Community Hospital (FBCH, Fort Belvoir, VA). Subjects of all race and gender seeking treatment for acute illness at these military hospitals were offered enrollment into the ARIC, IDCRP-045 and EPICC, IDCRP-085 protocols. EPICC study enrollment included subjects with laboratory-confirmed SARS-CoV-2 infection by nucleic acid amplification test, subjects with compatible illness in whom SARS-CoV-2 infection is initially suspected but PCR confirmed as SARS-CoV-2 negative, and asymptomatic subjects at risk of SARS-CoV-2 due to high risk exposure. Additionally, serum samples from 35 subjects undergoing treatment at JMS under the CAMP-NYC protocol were included in the assessment of assay performance. ARIC (IDCRP-045), EPICC (IDCRP-085) and CAMP-NYC protocols were approved by the Uniformed Services University Institutional Review Board.

### Multiplex microsphere-based immunoassay screening procedures

Three antigen-distinct MMIA were established: a) β-CoV MMIA that included SARS-CoV-2 spike and RBD, and SARS-CoV, MERS-CoV, HCoV-HKU1 and HCoV-OC43 spike; b) a SARS-CoV-2 and HCoV MMIA (SARS-2/HCoV) that included SARS-CoV-2, HCoV-HKU1, HCoV-OC43, HCoV-229E and HCoV-NL63 spike; and c) a SARS-CoV-2 spike and NP MMIA (SARS-2 spike/NP) (Table 1).

**Table 1.** Antigen composition of each MMIA.

| MMIA variation | Virus | Antigen |
| --- | --- | --- |
| β-CoV | SARS-CoV-2 | spike |
|  |  | RBD |
|  | SARS-CoV | spike |
|  | MERS-CoV | spike |
|  | HCoV-HKU1 | spike |
|  | HCoV-OC43 | spike |
| SARS-2/HCoV | SARS-CoV-2 | spike |
|  | HCoV-HKU1 | spike |
|  | HCoV-OC43 | spike |
|  | HCoV-229E | spike |
|  | HCoV-NL63 | spike |
| SARS-2 spike/NP | SARS-CoV-2 | spike |
|  |  | NP |

Serum samples were collected from venipuncture in serum separator tubes, processed and stored at -80 °C in 250 µL aliquots until use. At-home blood collection was performed with Mitra Collection Kits (Neoteryx, Torrance, CA, USA). Microsampler tip dried blood spots (DBS) were placed into deep-96-well microtiter plates containing 400 µL of 1X PBS-T and eluted overnight at 4 °C with agitation, 300 rpm. The duplicate microsampler tip DBS eluents were combined to 800 µL, vortexed, aliquoted to 200 µL and stored at -80 °C until antibody testing. For each 96-well plate, a multiplex master mix of antigen-coupled microspheres was made by diluting 100 µL of each antigen-coupled microsphere working stock into 10 mL (1:100) 1XPBS without calcium and magnesium (Corning Inc., Corning, NY) (all mentions of PBS refer to solutions without calcium and magnesium), and 100 µL of this master mix were added to each well so that each well contained 1 µL (∼23 ng) of each antigen-coupled microsphere per well. Wells were washed with 1XPBS + 0.05% Tween20 + 0.02% sodium azide two times. One hundred microliters of each serum sample was added to each well. Serum samples were diluted within a class II type A2 biological safety cabinet (BSC) then subjected to thermal inactivation for 30 min at 60 °C. Human serum samples (1.25 µL) were diluted 1:400 in PBS and eluted DBS aliquots were diluted 1:10 in PBS, and tested in technical duplicate A and B plates. Controls on each duplicate plate included a PBS blank (wells: A1, B1, G12, H12) and SARS-CoV-2 PCR-confirmed positive (C1, F12) and negative (D1, E12) serum sample. SARS-CoV-2 qualified controls for established expected inter- and intra-plate variations.

Samples were incubated at room temperature for 45 minutes with agitation (900 rpm), and plates were washed three times by an automated plate washer. Secondary antibody (goat anti-human IgG cross-absorbed biotin-conjugated or goat anti-human IgM cross-absorbed biotin-conjugated; Thermo Fisher Scientific, Waltham, MA) was diluted 1:5000 in 1XPBS + 0.05% Tween20 (PBST) and 100 µL of each secondary was added to each well and incubated for 45 minutes with agitation, and plates were washed three times. Streptavidin-phycoerythrin (Bio-Rad) was diluted 1:1000 in PBST and 100 µL was then added to each well and incubated for 30 minutes with agitation, and plates were washed three times. Lastly, 100 µL of PBST was added to each well and plates were resuspended by agitation for 5 minutes. Plates were read on Bio-Plex 200 multiplexing systems (Bio-Rad) with PMT voltage setting to the High RP1 target and 100 bead count requirements. Antibody levels are reported as quantified median fluorescence intensity (MFI). The average MFI of the four PBS-blank wells on each plate were subtracted from the MFI of each sample well and MFI values for samples are reported as the PBS adjusted average from duplicate plates.

### Threshold cutoffs for SARS-CoV-2 antibody

To establish threshold cutoffs for SARS-CoV-2 spike protein-specific antibody reactivity, we tested 127 archival acute and convalescent human serum samples from ARIC. Acute and convalescent serum samples were collected within approximately three and twenty-eight days of symptom onset, respectively. We established a cut-off of three standard deviations above the mean (99.7% probability) MFI of archival HCoV PCR-confirmed convalescent serum samples (n= 43) to establish a positivity threshold for detection of SARS-CoV-2 spike protein reactive IgG and IgM antibodies. The remaining 84 archival serum samples were tested against this MFI threshold cutoff for SARS-CoV-2 reactivity. The 127 archival ARIC serum samples were tested in technical duplicates in three independent experiments (β-CoV MMIA) and two independent experiments (SARS-2/HCoV MMIA and SARS-2 spike/NP MMIA) to establish threshold cutoffs and specificity for SARS-CoV-2. A highly reactive HKU1 PCR+ convalescent serum sample to SARS-CoV-2 NP was excluded in the 99.7% threshold sacrificing specificity for sensitivity.

### Non-human primate sera

Archived sera were used from rhesus macaques inoculated with a total dose of 2.6×10^6^ TCID50 of SARS-CoV-2 via a combination of intranasal, intratracheal, oral and ocular inoculation routes (41). Serum samples were collected at dpi 0 (baseline) and 21. SARS-CoV-2 IgG antibody seroconversion was determined as a 4-fold increase in MFI compared to the baseline sera collection.

### Statistical analysis

Figures were generated and statistical analyses were performed in GraphPad Prism version 7.0. When comparing zoonotic and endemic spike-specific IgG levels, MFI values were log10-transformed, checked for normality and parametric unpaired t-tests were employed to compare EPICC and JMS cohort to ARIC; statistical analysis with one-way ANOVA and Holm-Sidak’s multiple comparison were also utilized. When normality was not met, non-parametric unpaired Mann Whitney tests were performed. The positive predictive value and negative predictive value were calculated with MedCalc statistical software. ROC analysis was conducted using R version 4.0.2.

## RESULTS

### Generation of threshold cutoffs for SARS-CoV-2 antibodies

Despite low sequence similarity and identity between SARS-CoV-2 spike protein and seasonal HCoV spike proteins, pre-existing antibodies induced by prior infections with seasonal HCoVs are potentially cross-reactive, seemingly driven by conserved epitopes shared by the SARS-CoV-2 S glycoprotein S2 subunit (36, 37, 42). To enhance specificity for SARS-CoV-2 antibody detection, we established antibody positive/negative thresholds with SARS-CoV-2 naïve serum samples from human subjects with PCR-confirmed HCoV infections via the ARIC study (32, 40). First, we examined pre-existing IgG and IgM reactivity to SARS-CoV-2 antigens: spike, RBD and NP. IgG in convalescent sera collected ∼28 days after PCR-confirmed HCoV infections were more reactive with SARS-CoV-2 spike and NP than paired acute sera (Fig. 1A-C), suggesting that in a subset of persons recent HCoV-infection induces antibodies that are reactive with SARS-CoV-2 S glycoprotein and NP antigens. In some specific samples IgG in acute serum were more reactive with RBD (Fig. 1B).

**Figure 1.**
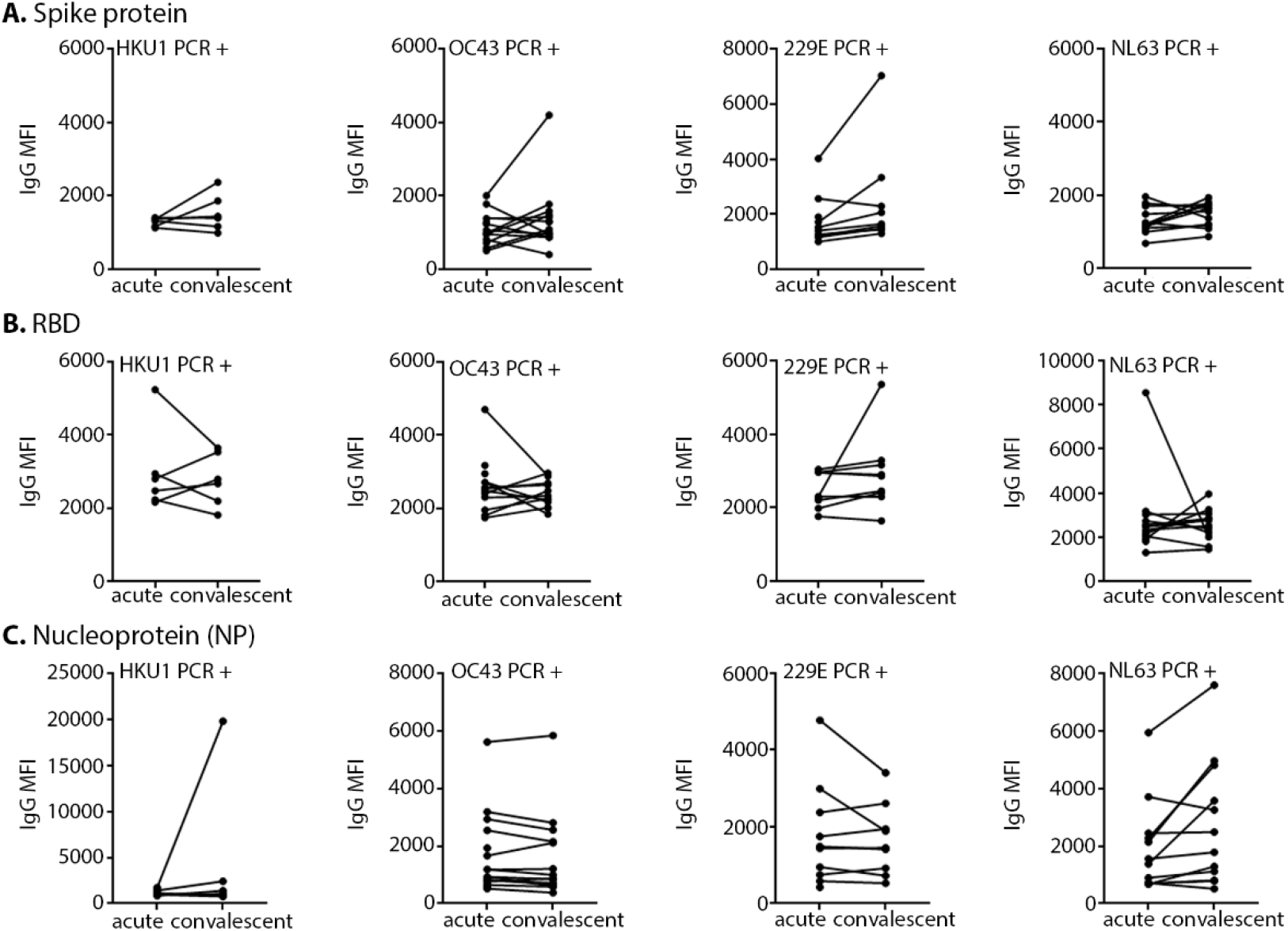
Pre-existing cross-reactivity with SARS-CoV-2 antigens informs threshold cutoffs for antibody positivity. Acute and convalescent serum samples from SARS-CoV-2 naïve HCoV PCR-positive subjects were tested in β-CoV **(A-B)** and SARS-CoV-2 spike/NP **(C)** MMIA. SARS-CoV-2 antigens are indicated. Subjects are grouped together based on HCoV PCR confirmation, HCoV-OC43 (n= 16), HCoV-HKU1 (n= 6), HCoV-NL63 (n= 13) and HCoV-229E (n= 10). MFI, median fluorescence intensity; IgG MFI values represent the MEAN of independent experiments performed in technical duplicates.

Second, accounting for this pre-existing cross-reactivity, we applied conventional probability distributions three standard deviations above the mean (99.7%) of these ARIC SARS-CoV-2 naïve HCoV-PCR confirmed convalescent sera to generate threshold cutoffs for SARS-CoV-2 positive antibodies (Fig. S1A-C). In addition to 99.7% probability distribution of ARIC sera, receiver operating characteristic (ROC) curve analysis of sera collected from SARS-CoV-2 RT-PCR confirmed subjects enrolled in the ongoing EPICC protocol were applied for spike, RBD and NP reactive IgG thresholds (Fig. S2A-D). ROC analyses identified MFI cutoff values, 4853 and 4144, retaining 100% sensitivity and 100% sensitivity with spike in both β-CoV and SARS-2/HCoV MMIA, respectively. When we applied this ROC curve value as a threshold for SARS-CoV-2 spike reactive IgG, we identified 2/43 (4.7%) serum samples from SARS-CoV-2 naïve ARIC with confirmed recent HCoV-OC43 and HCoV-229E infections above this cutoff (Fig. 1A). Further ROC analyses of antigen-specific IgG reactivity identified optimal MFI cutoff values for RBD of 4622 MFI (sensitivity= 82.7%, specificity= 91.5%) and NP of 6372 MFI (sensitivity= 90.5%, specificity= 100%). Pre-existing antibody reactivity with SARS-CoV-2 RBD and SARS-CoV-NP was further observed in 4/43 (9.3%) and 2/43 (4.7%), respectively, of ARIC serum samples from subjects with recent HCoV infections (Fig. 1B-C). These two distinct approaches to establish SARS-CoV-2 positive antibody threshold cutoffs resulted in very narrow positive/negative indeterminate ranges between the ARIC generated 99.7% threshold cutoff and the EPICC generated ROC curve threshold cutoff for spike, RBD and NP reactive IgG (Table 2).

**Table 2.** MFI threshold cutoffs for SARS-CoV-2 positive IgG.

| MMIA variation | antigen | 99.7% (ARIC) | ROC (EPICC) |
| --- | --- | --- | --- |
| β-CoV | spike | 4910 | 4853 |
|  | RBD | 4666 | 4622 |
| SARS-2/HCoV | spike | 4774 | 4144 |
| SARS-2 spike/NP | NP | 6649 | 6372 |

### Validation of multiplex microsphere-based immunoassay strategies

Next, we examined the influence of multiple antigen targets and multiplex composition on the diagnostic specificity and sensitivity of the MMIA strategies. Measurements of MMIA specificities were made using ARIC human serum samples, representing HCoV PCR-positive acute sera, rhinovirus PCR-positive acute/convalescent sera, and acute/convalescent sera from ‘no pathogen detected’ subjects, and were tested against our established threshold cutoffs. For sensitivity measurements, we utilized sera from SARS-CoV-2 PCR-confirmed human participants in EPICC and CAMP-NYC. Serum samples collected 10 – 73 dpso, median 23.5 dpso (IQR= 9.25) from hospitalized patients (n=35) at the JMS who participated in the cross-sectional CAMP-NYC protocol were screened for SARS-CoV-2 IgG antibody reactivity with MMIA strategies.

In the β-CoV and SARS-2 S-2P/NP MMIA strategies, serum samples (n= 116) collected 10 – 60 days post-symptom onset (dpso), median 37 dpso (IQR= 13.75), from SARS-CoV-2 PCR-positive outpatient (n= 62) and hospitalized patient (n= 54) participants enrolled in the EPICC protocol were tested for IgG reactivity with SARS-CoV-2 spike and RBD. Using the established threshold cutoffs, SARS-CoV-2 S-2P reactive IgG antibodies were only detected in PCR positive subjects from EPICC and JMS, retaining 100% specificity (Fig. 2A). The respective geometric mean IgG level with 95% confidence intervals (CI) against spike and RBD were 20,542 MFI (18,806-22,440 CI) and 18,271 MFI (16,251-20,542 CI) for EPICC samples (Fig. 2A-B), whereas, the geometric mean IgG level against spike and RBD were 26,829 MFI (25,757-27,945 CI) and 25,869 MFI (24,670-27,126) for JMS samples (Fig. 2A-B). These differences in geometric mean IgG levels are likely a reflection of COVID-19 severity as all JMS participants were hospitalized while outpatients made up a majority of the EPICC participants. Simultaneously utilizing spike and RBD proteins to capture target SARS-CoV-2 reactive IgG, we observed a significant correlation and strong linear relationship (Spearman r value= 0.9748; *P=* < 0.0001) between antibody binding to spike and RBD (Fig. 2C). SARS-CoV-2 spike reactive IgG antibody detection by β-CoV MMIA was calculated with 95% CI as follows, sensitivity= 99.3% (96.3%-99.9% CI), specificity= 100% (95.7%-100.0% CI). Assuming a US disease prevalence of 5.0%, the the β-CoV MMIA had a positive predictive value (PPV) = 100.0%, and negative predictive value (NPV) = 99.9% (99.8%-100.00% CI) (Table 3). Comparatively, utility of SARS-CoV-2 RBD for IgG detection had a slightly reduced sensitivity= 96.0% (91.5%-98.5% CI), specificity= 96.4% (89.9%-99.2% CI), PPV= 58.6% (31.8%-81.1% CI) and NPV= 99.8% (99.5%-99.9% CI) with a 5.0% prevalence estimate.

**Table 3.** β-CoV MMIA performance.

|  | SARS-CoV-2 PCR Status/Archival Sera |  |  |  |
| --- | --- | --- | --- | --- |
| SARS-CoV-2<br>spike IgG<br>Antibody Test |  | Positive<sup>1</sup> | Negative | Total |
|  | Positive | 149 | 0 | 149 |
|  | Negative<sup>2</sup> | 1 | 84 | 85 |
|  | Total | 150 | 84 | 234 |
|  | Sensitivity | 99.33 |  |  |
|  | Specificity | 100% |  |  |
| SARS-CoV-2<br>RBD IgG<br>Antibody Test |  | Positive | Negative | Total |
|  | Positive | 144 | 3 | 147 |
|  | Negative | 6 | 81 | 87 |
|  | Total | 150 | 84 | 234 |
|  | Sensitivity | 96.00% |  |  |
|  | Specificity | 96.43% |  |  |

<sup>1</sup>Performance analysis included serum samples from n= 116 PCR positive subjects including 62 outpatients, 54 hospitalized subjects and 34 JMS hospitalized subjects
<sup>2</sup>Negative samples were drawn from the pre-2019 ARIC sera collection

**Figure 2.**
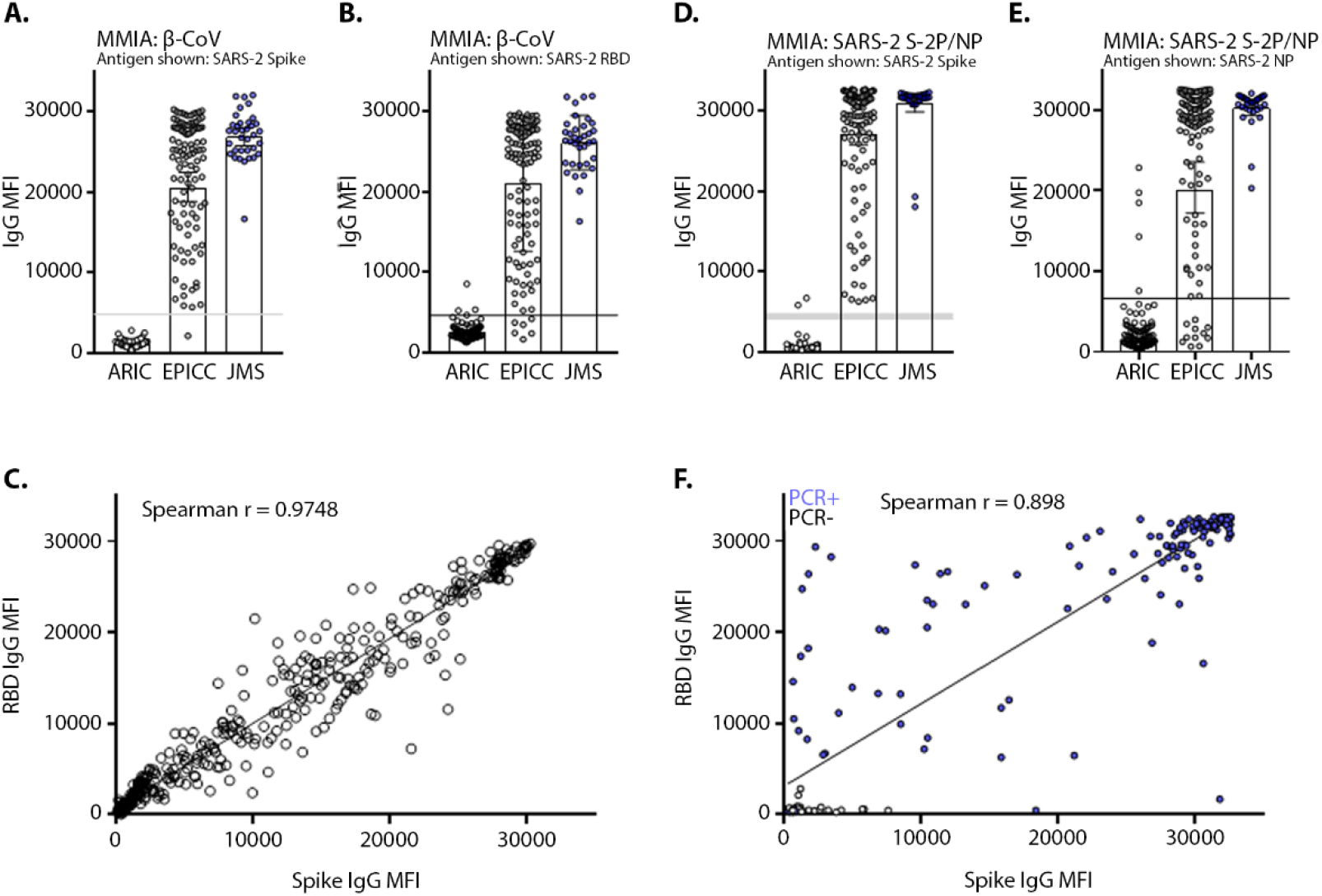
Detection of SARS-CoV-2 reactive serum IgG with S glycoprotein and nucleoprotein antigens. Serum samples were tested in β-CoV MMIA for reactivity with SARS-CoV-2 spike **(A)** and RBD **(B). (C)** Correlation analysis of IgG binding to SARS-CoV-2 spike and RBD when tested simultaneously in the β-CoV MMIA. Serum samples were tested in SARS-2 spike/NP MMIA for reactivity with SARS-CoV-2 spike **(D)** and NP **(E). (F)** Correlation analysis of IgG binding to SARS-CoV-2 spike and NP when tested simultaneously in the SARS-2 spike/NP MMIA.

Utility of a SARS-CoV-2 NP antigen was explored in a SARS-CoV-2 spike/NP MMIA able to simultaneously capture antibodies specific to both S glycoprotein and NP. The respective, geometric mean IgG levels of spike and NP were 25,526 MFI (23,703-27,490 CI) and 20,165 MFI (17,241-23,584 CI) in EPICC, and 30,723 MFI (29,434-32,069) and 30,268 MFI (29,350-31,215 CI) in JMS sera. In the absence of HCoV spike, two ARIC serum samples were cross-reactive with SARS-CoV-2 spike above the positive threshold, and all SARS-CoV-2 PCR-confirmed samples had detectable spike reactive IgG (Fig. 2D). Five ARIC samples were cross-reactive with SARS-CoV-2 NP and 11 EPICC subjects developed IgG to spike but not NP (Fig. 2E). Regardless, the correlation between spike and NP was statistically significant (Spearman r value= 0.898; *P=* < 0.0001) (Fig. 2F). Antigen-specific immune responses to spike and NP have been observed in COVID-19 patients and are associated with disease severity and older age (4, 43, 44). The performance of SARS-CoV-2 NP IgG assay sensitivity= 92.7% (87.3%-96.3% CI), specificity= 94.1% (86.7%-98.0% CI), PPV= 45.0% (25.9%-65.8% CI) and NPV= 99.6% (99.3%-99.8%) (Table 4).

**Table 4.** SARS-2 spike/NP MMIA performance.

| SARS-CoV-2<br>NP IgG<br>Antibody Test | SARS-CoV-2 PCR Status/Archival Sera |  |  |  |
| --- | --- | --- | --- | --- |
|  |  | Positive | Negative | Total |
|  | Positive | 139 | 5 | 144 |
|  | Negative | 11 | 79 | 90 |
|  | Total | 150 | 84 | 234 |
|  | Sensitivity | 92.7% |  |  |
|  | Specificity | 94.1% |  |  |

SARS-CoV-2 IgG seroconversion has been detected early after exposure and sometimes in parallel with IgM seroconversion (45-47). For SARS-CoV-2 IgM detection, again we applied a ROC analysis threshold, 1445 MFI (Fig. S3A), which was more conservative than the 99.7% probability threshold cutoff, 840 MFI (Fig. S3B). No ARIC sera were reactive above the threshold for SARS-CoV-2 spike protein IgM, and EPICC spike geometric IgM levels were 2612 MFI (1684-2775 95% CI) and JMS spike geometric IgM levels were 11,595 MFI (9438-14,244 95% CI) (Fig. S3C). EPICC sera tested represent a cohort of 62% outpatients, and differences in detectable IgM levels between EPICC and JMS are probably reflections of COVID-19 clinical outcomes that separate cohort. The SARS-CoV-2 spike reactive IgM detection sensitivity was lower than IgG, with performance analysis conducted with serum samples collected ≥ 7 days post-symptom onset, sensitivity= 73.33% (65.51%-80.22% CI), specificity= 100.00% (95.70-100.00%), PPV= 100.00 and NPV= 98.62% (98.20%-98.95%) (Table S1).The temporal window to capture SARS-CoV-2 IgM was shorter than IgG, and as serum samples included in β-CoV MMIA performance analysis ranged from 10 – 60 dpso, lower IgM sensitivity is further driven by outpatient enrollments in the EPICC protocol with an average 28 dpso that were IgG positive, but IgM negative (Table S2).

### SARS-2/HCoV spike MMIA

The interplay of pre-existing HCoV spike-specific antibodies and COVID-19 outcomes, as well as post vaccine responses, remains a critical question. As such, we also validated a SARS-2/HCoV spike MMIA strategy that can simultaneously measure antibodies to SARS-CoV-2 and all four seasonal HCoVs. To validate this MMIA strategy, we utilized EPICC serum samples (n= 148) from SARS-CoV-2 PCR-positive participants collected 7 – 60 dpso (median= 35, IQR= 23). Again, we detected no SARS-CoV-2 S-2P IgG positives in ARIC sera, and geometric mean IgG levels were 21,487 MFI (19,568-23,584 CI) and 30,871 MFI (30,403-31,346 CI) from EPICC and JMS serum samples, respectively (Fig. 3A). In addition to serum samples collected at hospitals, the EPICC protocol provides at-home blood collection of capillary blood samples as DBS for longitudinal serology. Of additional importance, we validated paired DBS IgG against serum collected by serum separator tubes (SST) and observed a significant and strong linear correlation of SARS-CoV-2 MFI derived from DBS and SST collected blood specimens (Fig. 3B). Reports have demonstrated anti-S glycoprotein IgG seroconversion between 3 and 14 dpi (41, 48-50), to further assess the MMIA strategy, sensitivity for the SARS-2/HCoV MMIA was measured within an early and narrow range of EPICC and JMS blood specimen collection, 7-28 dpso. SARS-2/HCoV spike MMIA assay performance was, sensitivity= 94.4% (86.4%-98.5% CI), specificity= 100.0% (95.7%-100.0% CI), PPV= 100.0% and NPV= 99.7% (99.3%-99.9% CI) (Table 5). As previously seen with the β-CoV assay, inclusion of HCoV spike improves specificity for SARS-CoV-2; both β-CoV and SARS-2/HCoV MMIAs have near identical upper threshold cutoff values, 4910 MFI and 4774 MFI, thus the inclusion of HCoV-HKU1 and HCoV-OC43 spike led to improvements in specificity.

**Table 5.** SARS-2/HCoV MMIA performance.

| SARS-CoV-2<br>spike IgG<br>Antibody Test | SARS-CoV-2 PCR Status/Archival Sera |  |  |  |
| --- | --- | --- | --- | --- |
|  |  | Positive | Negative | Total |
|  | Positive | 68 | 0 | 68 |
|  | Negative | 4 | 84 | 88 |
|  | Total | 72 | 84 | 156 |
|  | Sensitivity | 94.4% |  |  |
|  | Specificity | 100% |  |  |

<sup>1</sup>IgG antibody test included serum samples collected 7-28 days post-onset from PCR positive EPICC and JMS participants.

**Figure 3.**
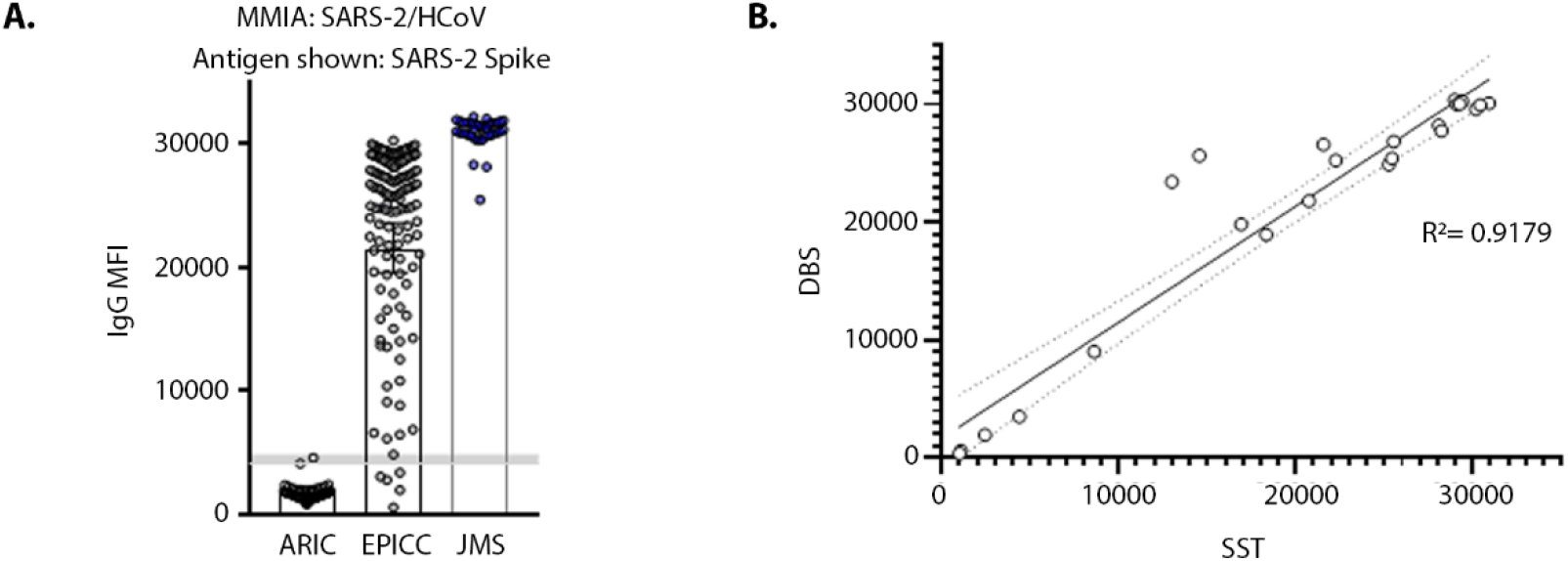
SARS-2/HCoV MMIA detection of SARS-CoV-2 IgG in blood specimens. **(A)** Serum samples were screened with a SARS-2/HCoV spike protein MMIA and reactivity to SARS-CoV-2 spike is shown; shaded grey bar indicates the threshold cutoff for IgG positivity. **(B)** Paired blood specimens (n= 22) collected from capillary blood as a dried blood spot (DBS) and serum collected by serum separator tubes (SST) were tested with the SARS-2/HCoV spike protein MMIA; SARS-CoV-2 spike reactive IgG MFI is indicated on x- and y-axes.

### COVID-19 subject antibody reactivity with zoonotic and endemic coronaviruses

Next, we investigated SARS-CoV-2 *de novo* IgG antibody cross-reactivity with SARS-CoV and MERS-CoV spike proteins. Threshold cutoffs for SARS-CoV and MERS-CoV spike reactive IgG were similarly set with ARIC sera as detailed for SARS-CoV-2 spike (Fig. 4A). An indeterminate range (3840 – 4910 MFI) for positive/negative IgG reactivity was established with SARS-CoV-2, SARS-CoV and MERS-CoV spike protein reactive IgG MFI and 4910 MFI represents the threshold for positivity. We compared levels of SARS-CoV and MERS-CoV reactive IgG in SARS-CoV-2 PCR positive subjects. SARS-CoV and MERS-CoV S glycoprotein share 82% and 50% homology, respectively, with SARS-CoV-2 S glycoprotein (37). A significant level of cross-reactive antibodies to SARS-CoV and MERS-CoV were observed in SARS-CoV-2 PCR positive and IgG positive EPICC and JMS cohort sera, but not SARS-CoV-2 naïve ARIC sera (Fig. 4B). The geometric mean IgG MFI levels of SARS-CoV reactive IgG in EPICC and JMS were 4007 (3422 – 4693 CI) and 9307 (7423 – 11,669 CI), respectively; and the geometric mean IgG MFI levels of MERS-CoV reactive IgG in EPICC and JMS were 4255 (3517 – 5173 CI) and 8307 (6436 – 10,722 CI), respectively.

**Figure 4.**
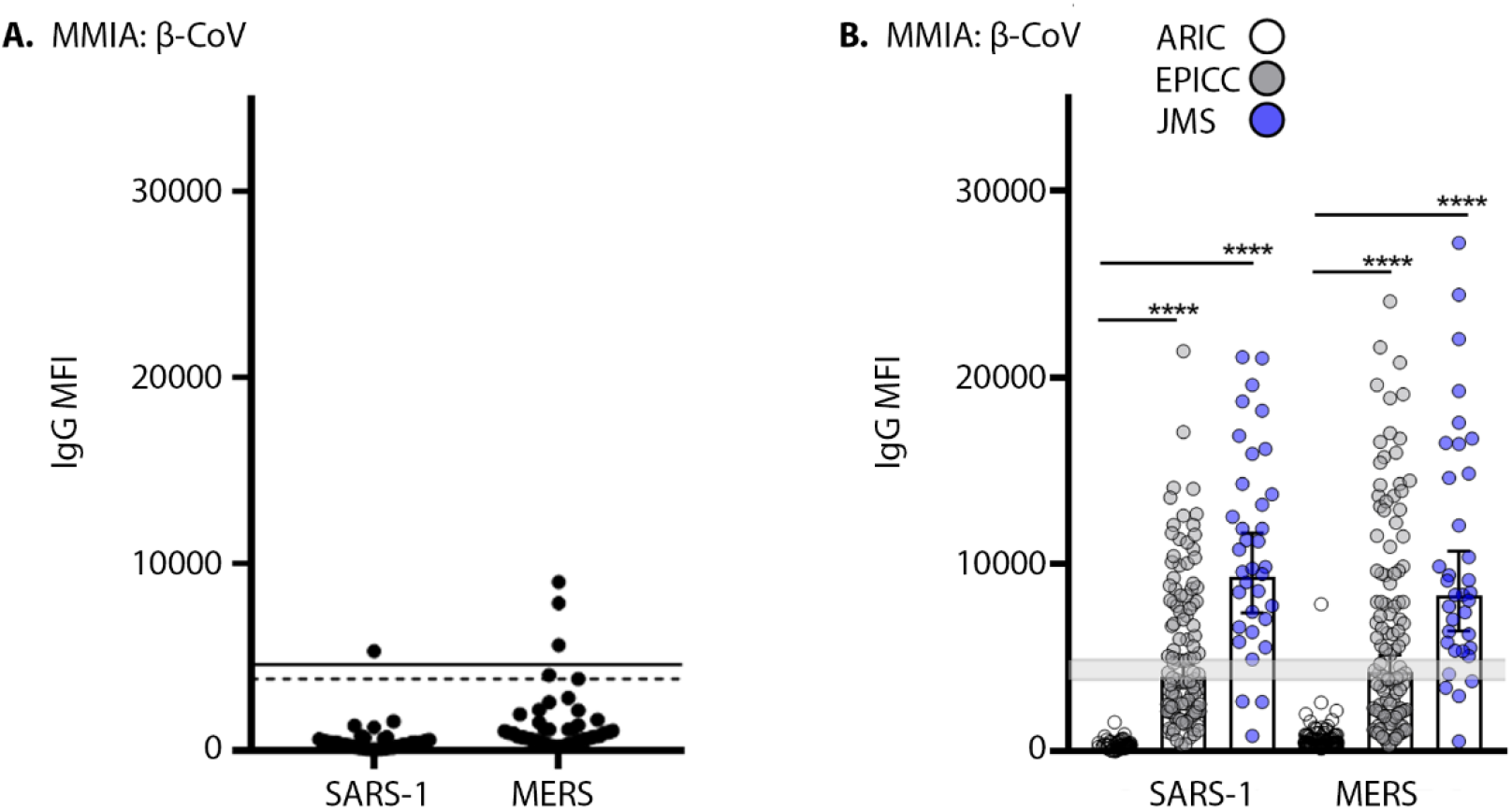
SARS-CoV and MERS-CoV IgG increases after SARS-CoV-2 infection. **(A)** 99.7% probability distribution of ARIC HCoV PCR+ convalescent serum samples (n= 43) reactive with SARS-CoV (SARS-1) and MERS-CoV (MERS) spike; a dashed line and solid line indicates the SARS-CoV and MERS-CoV spike protein threshold cutoff values, respectively. **(B)** Sera from SARS-CoV-2 positive EPICC and JMS cohorts were tested in a β-CoV MMIA for IgG reactivity to SARS-CoV and MERS-CoV spike, a shaded grey line indicates the threshold cutoff for IgG positivity; error bars indicate the geometric mean and 95% confidence intervals; unpaired Mann-Whitney t-tests of EPICC and JMS compared to ARIC, **** *P*-values= < 0.0001.

In the absence of HCoV naïve human cohort, we turned to SARS-CoV-2 non-human primates (NHP) model to inform the stimulation of HCoV cross-reactive antibodies. NHP had no evidence of *de novo* IgG reactivity with HCoV-HKU1 and HCoV-OC43 spike proteins after SARS-CoV-2 challenge and seroconversion (Fig. 5A). We extrapolated the 99.7% indeterminate range of SARS-CoV, SARS-CoV-2 and MERS-CoV spike protein reactive IgG as a cutoff for HCoV reactive antibodies (Fig. 4A). We found that IgG reactivity with HCoV spike proteins in the SARS-CoV-2 PCR positive patient serum samples were significantly higher than those in SARS-CoV-2 naïve ARIC cohort. We found significant geometric mean IgG increases of HCoV-OC43, 26,518 (25,920-27,131 CI) and 29,454 (28,759-30,167 CI) in EPICC and JMS cohorts, respectively; HCoV-HKU1 geometric mean IgG levels in EPICC and JMS cohorts were 15,739 (14,693-16,859 CI) and 19,360 (17,027-22,012 CI), respectively (Fig. 5B). Furthermore, we detected a significant increase in IgG-reactive with HCoV-NL63 associated with SARS-CoV-2 infection in EPICC and JMS cohorts; a significant increase in HCoV-229E IgG was detected in JMS sera (Figure 5C).

**Figure 5.**
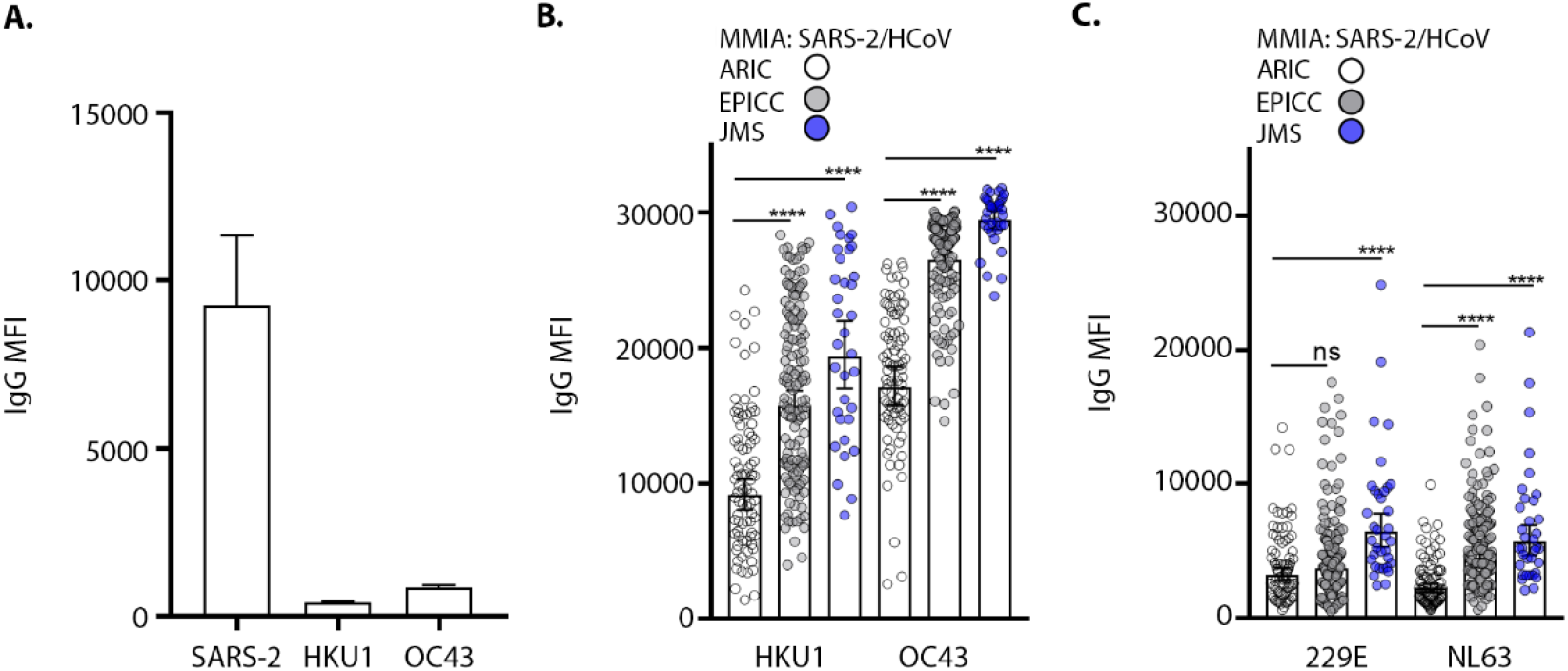
SARS-CoV-2 infection is associated with rises in HCoV antibody levels. **(A)** Serum samples collected 21 dpi from SARS-CoV-2 challenged non-human primates (n= 4) and tested in two independent experiments performed in technical duplicates with a SARS-2/HCoV spike protein MMIA; error bars represent mean±SD. IgG reactivity with seasonal HCoV, -HKU1 and -OC43 (β-CoVs) **(B)** and -229E and -NL63 (α-CoVs) **(C)**, spike proteins were tested with a SARS-2/HCoV spike MMIA. Error bars indicate the geometric mean and 95% CI, IgG levels were compared by unpaired t-tests with Welch’s correction of EPICC and JMS compared to ARIC, **** *P*-values= < 0.0001.

## DISCUSSION

In this study, we have demonstrated that use of a multiplex microsphere-based immunoassay (MMIA) built using Luminex xMAP-based technology in which individual microspheres are bound to pre-fusion stabilized S glycoprotein trimers of SARS-CoV-2 and the seasonal endemic HCoVs enables sensitive and specific detection of SARS-CoV-2 IgG antibodies. Our SARS-CoV-2 spike-based MMIA strategies have sensitivities ranging 94 – 99 % as early as 7 – 10 days after symptom onset in PCR-confirmed cases of SARS-CoV-2 infection and 100% specificity for SARS-CoV-2 IgG, comparable with several other EUA serology tests (6). Conserved epitopes present in the prefusion stabilized native-like trimeric S glycoprotein oligomers (spike protein) are the likely major factor in the observed cross reactions between the coronavirus S glycoproteins, affecting specificity for some serology assays. By using HCoV-HKU1 and HCoV-OC43 spike proteins to capture pre-existing antibodies that would be cross-reactive with SARS-CoV-2 spike, the assay had a 100% specificity for SARS-CoV-2 serology. Importantly, the ability to simultaneously capture SARS-CoV-2 spike and NP-specific antibodies within a single assay will facilitate high-throughput approaches for differentiating antibody responses between SARS-CoV-2 natural infections and vaccinations.

The magnitude of the antibody response to SARS-CoV-2 infection has been associated with COVID-19 severity (51).This was apparent in our validation tests as geometric mean IgG levels were consistently elevated in sera from the JMS cohort, comprised of all hospitalized patients, whereas sera from the EPICC cohort had lower geometric mean IgG to SARS-CoV-2 antigens, of which the majority of EPICC study participants were outpatients. Interestingly, geometric mean IgG level, 20,542 MFI (18,806-22,440 CI) to SARS-CoV-2 spike determined by testing with the β-CoV MMIA was lower than the geometric mean IgG levels spike, 25,226 MFI (23,703-27,490 CI) in the SARS-2 spike/NP MMIA. One possibility is that in the β-CoV MMIA, inclusion of spike and RBD which represent overlapping epitopes leads to competition for antibodies, decreasing overall MFI levels to either S glycoprotein antigen individually. The effect of this binding competition could negatively affect durability studies of longitudinal samples when concentrations of circulating SARS-CoV-2 sera IgG begins to wane. Whereas establishing the SARS-CoV-2 spike protein and NP MMIA will have utility for the future differentiation between antibody responses to SARS-CoV-2 vaccination and natural infection induced antibody responses, with some inherent limitations to sensitivity and specificity.

Of additional importance, the MMIA approach demonstrated *de novo* IgG cross-reactivity with SARS-CoV and MERS-CoV in the SARS-CoV-2 PCR+/IgG+ EPICC and JMS cohorts compared with archival sera (ARIC). The EPICC geometric mean IgG levels reactive with SARS-CoV spike were near the indeterminate range of positivity, suggesting that significant rises in IgG levels after SARS-CoV-2 are driven by a subset of subjects that develop specific B cell repertoires that can be cross-reactive with SARS-CoV. A higher geometric mean IgG level to SARS-CoV and MERS-CoV were detected in JMS sera, which includes all hospitalized patients and suggests that induction of cross-reactivity is associated with COVID-19 severity. Conserved cross-neutralizing epitopes between SARS-CoV and SARS-CoV-2 S glycoproteins have been identified (52, 53), whether SARS-CoV-2 induced *de novo* IgG antibody responses to SARS-CoV and MERS-CoV spike proteins detected with this MMIA strategy are retained after affinity maturation, or are cross-neutralizing requires further investigation. The induction of cross-reactive SARS-CoV, SARS-CoV-2 and MERS-CoV antibodies further demonstrates that shared spike proteins epitopes exist and that rational-vaccine designs may be able to develop pan-zoonotic coronavirus vaccines.

Here, we observed increases in seasonal HCoV spike reactive IgG antibodies in SARS-CoV-2 positive cohort. SARS-CoV-2 has been shown to stimulate OC43 memory B cells through conserved epitopes in the SARS-CoV-2 S glycoprotein S2 subunit and serum samples from subjects with recent HCoV infection contain SARS-CoV-2 cross-reactive but not cross-neutralizing antibodies (54). As *de novo* IgG cross-reactivity with HCoV-HKU1 and HCoV-OC43 spike protein was not observed in SARS-CoV-2 challenged NHPs (Figure 5A), the presence of prior humoral memory to seasonal HCoV appears necessary to drive cross-reactivity. Overall, the OC43 S glycoprotein only shares 30 to 40% amino acid sequence identity/similarity with SARS-CoV-2 S glycoprotein (37). The S1 subunit, wherein resides the RBD, has more sequence variance among OC43 and SARS-CoV-2, in contrast to the S2 subunit heptad repeat regions where amino acid sequence similarity is between 50 to 75%. It would seem unlikely that a SARS-CoV-2 *de novo* IgG response would result in cross-reactive antibodies that would bind at immunoassay saturation to distantly-related seasonal β-CoVs. However, we did observe increases in HCoV-NL63 spike reactive antibodies EPICC and JMS cohorts, suggestive that SARS-CoV-2 infection and the subsequent heightened inflammatory state can induce a humoral response with enough antibody diversity capable of binding to distantly-related human α-CoVs. It will be important to expand on this observation with larger numbers of subjects in different age groups to evaluate the extent of SARS-CoV-2 stimulated seasonal HCoV memory responses.

Other, preliminary evidence has estimated a negative relationship between HKU1 and OC43 back-boosted responses and the generation of SARS-CoV-2 neutralizing antibodies (55), implying that in some subjects OC43 immune imprinting may have similar effects akin to immunological imprinting, antigenic epitope masking and/or *original antigenic sin* (56, 57). Whether HCoV immune imprinting has any association with clinical outcomes or SARS-CoV-2 antibody longevity requires further investigation. Back-boosting stimulation of a cross-reactive memory response might also explain cases of synchronous SARS-CoV-2 IgM and IgG seroconversion and IgG seroconversion prior to IgM (58). To our knowledge there is no evidence that HCoV-induced antibodies promote clinical protection in SARS-CoV-2 infected individuals. The vast majority of the SARS-CoV-2 confirmed participants in these studies had high levels of HCoV-OC43 IgG; however, they were also seeking treatment for mild to moderate/severe COVID-19 which requires further investigation. Larger, prospective, longitudinal observational studies in which serum samples are obtained before infection may ultimately be required to definitively determine if HCoV-induced antibodies confer any protection against COVID-19 and if the presence of HCoV memory affects the longevity and development of a protective SARS-CoV-2 humoral response.

## Data Availability

The data that support the findings of this study are available from the corresponding author upon reasonable request.

## DECLARATIONS

These research protocols, IDCRP-085, IDCRP-045 and CAMP-NYC, were approved by the USU IRB. The data that support the findings of this study are available from the corresponding author(s upon reasonable request.

## ETHICS STATEMENT

The referenced human subjects protocols (IDCRP-045, IDCRP-085, and CAMP-NYC) were approved by the Uniformed Services University Institutional Review Board and participating sites. All subjects provided written or verbal informed consent using approved documents and procedures; the consent forms include clauses allowing use of specimens for investigations including those conducted in this study.

## DISCLAIMER

The contents of this publication are the sole responsibility of the author(s) and do not necessarily reflect the views, opinions, or policies of the Uniformed Services University (USU), the Henry M. Jackson Foundation for the Advancement of Military Medicine, Inc. (HJF), National Institutes of Health or the Department of Health and Human Services, Brooke Army Medical Center, the U.S. Army Medical Department, the U.S. Army Office of the Surgeon General, the US Department of Defense (DoD), the Departments of the Air Force, Army or Navy, or the U.S. Government. Mention of trade names, commercial products, or organization does not imply endorsement by the U.S. Government. A number of the co-authors are military service members (or employees of the U.S. Government). This work was prepared as part of their official duties. Title 17 U.S.C. §105 provides that ‘Copyright protection under this title is not available for any work of the United States Government.’ Title 17 U.S.C. §101 defines a U.S. Government work as a work prepared by a military service member or employee of the U.S. Government as part of that person’s official duties.

## ACKNOWLEDGEMENTS

We thank Kelly Snead, Vanessa Wall, John-Paul Denson, Simon Messing, and William Gillette (Protein Expression Lab, FNCLR) for excellent technical assistance. We also thank Kathleen Pratt (Department of Medicine, USUHS) for assistance with CAMP-NYC sample acquisition.

