## Supplementary Appendix for "Antigen-based multiplex strategies to discriminate SARS-CoV-2 natural and vaccine induced immunity from seasonal human coronavirus humoral responses"

**Table S1. β-CoV MMIA performance**

| | SARS-CoV-2 PCR Status/Archival Sera | | | |
|---|---|---|---|---|
| SARS-CoV-2 spike IgM Antibody Test | | Positive | Negative | Total |
| | Positive | 110 | 0 | 110 |
| | Negative | 40 | 84 | 124 |
| | Total | 150 | 84 | 234 |
| | Sensitivity | 73.33% | | |
| | Specificity | | 100.00% | |

**Table S2. IgG and IgM seropositivity within 28 days post-symptom onset (dpso)**

| dspo | IgG+ | IgG+/IgM+ |
|---|---|---|
| 7 – 14 | 80.0% (12/15) | 73.3% (11/15) |
| 15 – 28 | 100% (31/31) | 93.5% (29/31) |

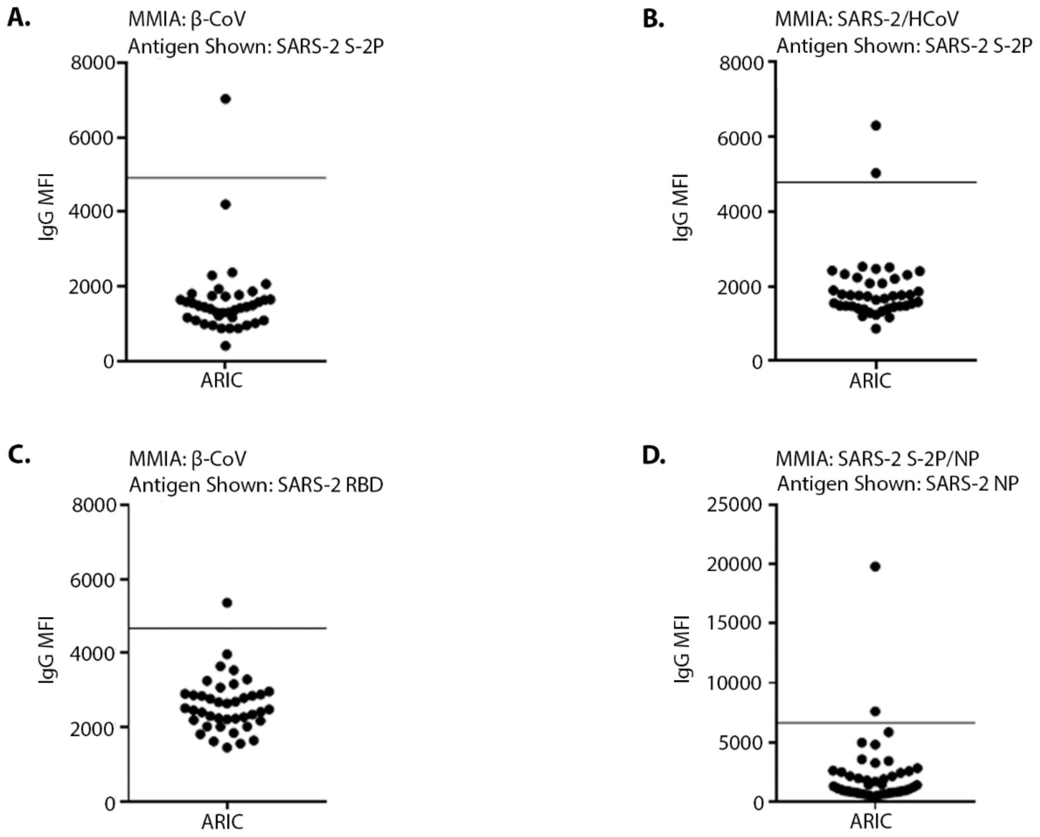


**Figure S1. 99.7% probability distribution threshold cutoffs for SARS-CoV-2 antigen reactive IgG.** Convalescent serum samples (n=43) from SARS-CoV-2 naïve ARIC subjects with HCoV PCR-confirmed infections were tested with MMIA indicated in panels. Antigens shown as follows: **(A)** SARS-CoV-2 spike (S-2P), **(B)** SARS-CoV-2 spike (S-2P), **(C)** SARS-CoV-2 RBD, and **(D)** SARS-CoV-2 NP; sold lines indicate the mean+3SD (99.7%) MFI.

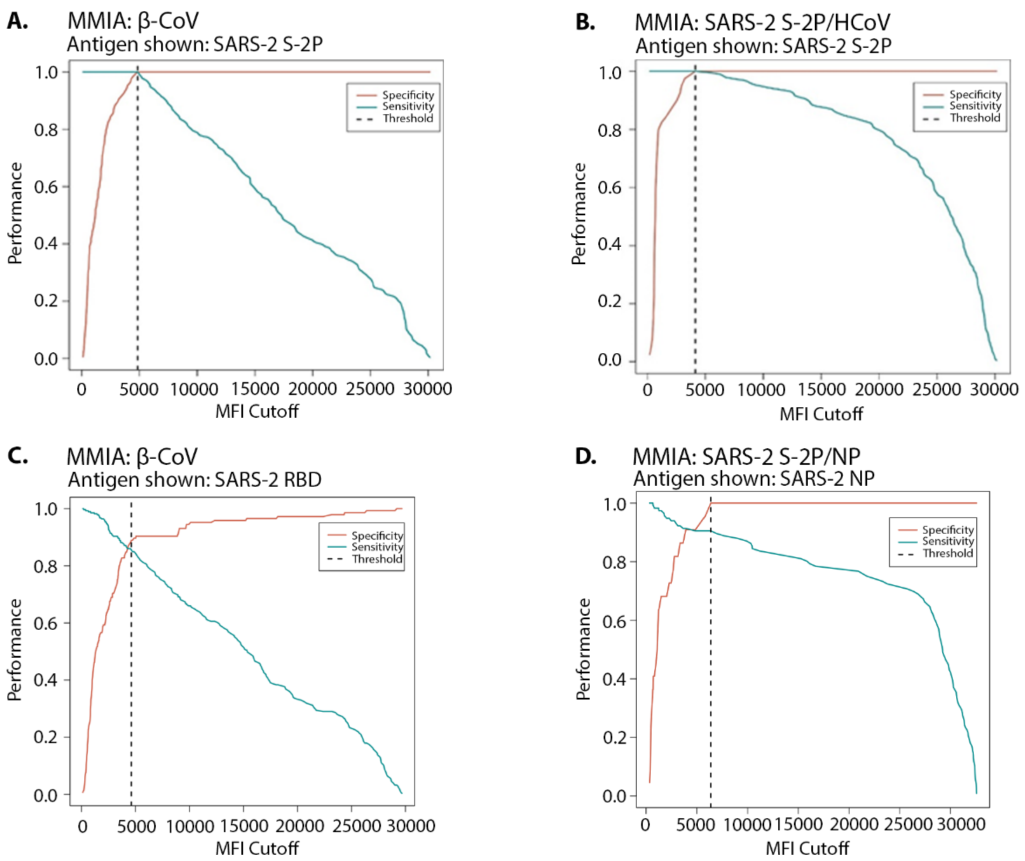


**Figure S2. Receiver operating characteristic (ROC) curve analysis of SARS-CoV-2 PCR-positive and -negative serum samples from the EPICC study provided a second measurement of threshold cutoffs for SARS-CoV-2 antigen reactive IgG.** EPICC Sera were tested with MMIA indicated in panels; antigens shown as follows: **(A)** SARS-CoV-2 spike (S-2P), **(B)** SARS-CoV-2 spike (S-2P), **(C)** SARS-CoV-2 RBD, and **(D)** SARS-CoV-2 N

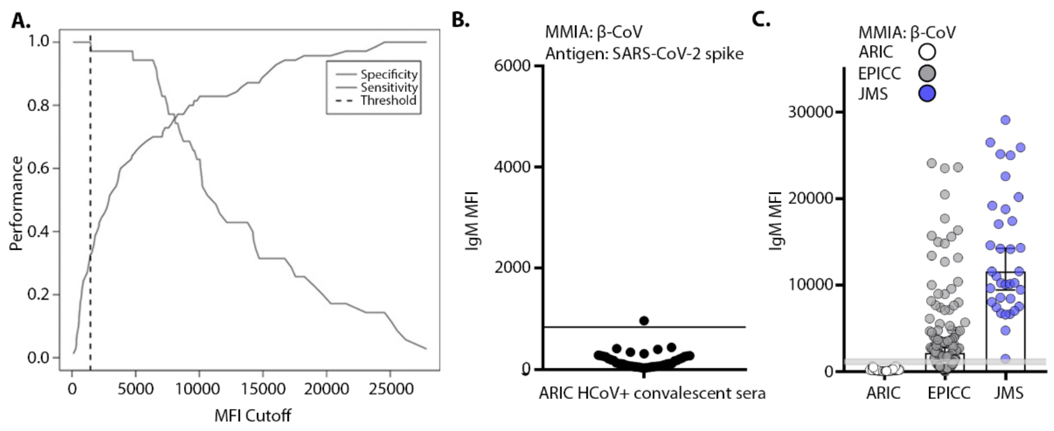


**Figure S3. SARS-CoV-2 spike reactive IgM detection with the β-CoV MMIA.**

**(A)** ROC curve analysis of SARS-CoV-2 spike protein IgM antibody reactivity in a β-CoV MMIA. PCR-confirmed SARS-CoV-2 positive and negative serum samples (n= 105) were tested and 100% specificity was achieved at threshold cutoff of 1446 MFI. **(B)** SARS-CoV-2 naïve ARIC HCoV PCR-positive convalescent serum samples (n=43) were tested to establish a 99.7% probability threshold (840 MFI). **(C)** ARIC (n=84), SARS-CoV-2 PCR-positive EPICC (n=116) and JMS (n=35) serum samples were tested in technical duplicates.
